# Can medication mentions in CRIS be used for researching medication use in older people with dementia? Comparing the natural language processing app for medicines to GP prescribing

**DOI:** 10.1101/2023.01.27.23285104

**Authors:** Katrina A.S. Davis, Matthew Broadbent, Delia Bishara, Christoph Mueller, Robert Stewart

**Affiliations:** KCL Institute of Psychiatry Psychology and Neuroscience, London, UK; South London and Maudsley NHS Foundation Trust, London, UK

## Abstract

**Background:** The efficacy of long-term preventative medication in people who have dementia and other comorbidities is unknown. Electronic health record-based observational studies may play a role in evaluating medicines, and SLaM-CRIS is one such resource. Medication in SLaM-CRIS is extracted from structured and unstructured fields using a natural language processing app. We aimed to compare the results from the medication app with GP prescribing, using an innovative data link with primary care (Lambeth DataNet, LDN) that covers around one-quarter of the SLaM-CRIS population.

**Methods:** A cohort was created of people with both LDN record and SLaM-CRIS record who had received a diagnosis of dementia in either record, in which ascertainment of medication could be compared. Ten classes of medication commonly taken long-term for preventative purposes were studied (aspirin, statins, ACE inhibitors, anticoagulants, beta-blockers, bisphosphonates, calcium with vitamin D, non-aspirin antiplatelets, proton pump inhibitors and antidepressants), plus medications taken for dementia itself. Mentions of these medication classes in SLaM-CRIS from around the time of dementia diagnosis were extracted using a natural language processing application. Prescription issue for the same medications was extracted from LDN in the year of dementia diagnosis and compared with that from SLaM-CRIS on a cohort and individual level.

**Results:** Our sample included 4410 with documentation of dementia in either SLaM-CRIS or LDN. Estimation of the prevalence of the use of each medication in CRIS was compared to LDN, and was within +/-3 per 100, except for calcium with vitamin D supplement, where SLaM-CRIS prevalence was 13 people per 100 lower than LDN (8.6% vs 21.2%). Medication ascertainment of all groups except calcium with vitamin D supplement showed good agreement (kappa above 0.7), and very good agreement for antidepressants and dementia drugs (kappa above 0.8). Sensitivity was highest for dementia drugs and antidepressants (above 90%), 85% for statins and 75% for aspirin. Restricting to those with a memory clinic referral did not change the levels of agreement.

**Discussion:** Routinely collected data cannot provide a gold-standard measure of what medications are truly taken by patients with dementia, but several sources can provide a proxy measure. This analysis supports the use of the natural language processing application for medication in SLaM-CRIS to extract medication mentions of relevance to people with dementia, as compared to prescribing from the GP at around the same time. However, some medications show low sensitivity, possibly due to low recording or inconsistency in the text used to record, and enhancement will be needed before studying these medications.

## Background

Older people with dementia often have other disorders or risk states for which medications can be prescribed. While much of this medication will be recommended in single-condition guidelines using evidence from trials of people with that condition only, it is not clear whether people with multimorbidity or frailty benefit in the same way.(Bishara and Harwood 2014, National Guideline Centre 2016, Mueller, Molokhia et al. 2018, Licher 2022, Longo, Burnett et al. 2022) It is therefore important to look at evidence specifically in people with dementia to inform clinicians and patients. Randomised-controlled trials are often unlikely, as they are expensive to run and burdensome for patients, which means that observational studies are more likely to provide an indication as to the risks and benefits of treatment.(Davis, Farooq et al. 2019)

Electronic health records (EHR) repositories or databases can provide data for observational studies, by repurposing medical information with governance to protect confidentiality. Separate health records, and EHRs more recently, exist for each organisation (such as GP practice) or group of organisations (such as an NHS Trust with multiple hospitals), containing patient-centred data for use by clinicians and administrators in that entity. This can mean that people interacting with multiple organisations may have data in many EHRs, some of which may be available for research. In South East London in the UK, someone who saw their GP, was referred to the mental health service memory clinic, and also was seen in the A&E for chest pain would have a record in the EHRs of the GP practice, mental health trust and general hospital.

Information in EHRs, entered mainly by administrators and clinicians, has two types – structured data and unstructured text. Structured data will be things like age and sex, alongside diagnosis codes and prescription records. Unstructured text may contain much of the contextual information such as uncertainties around a diagnosis, why a prescription was written and whether it was working. When EHR data is repurposed, it is common for researchers to only access structured data. Text may be more difficult to use in large-scale studies and also has confidentiality issues.

The South London and Maudsley NHS Foundation Trust (SLaM) provides specialist mental health and dementia services for four boroughs in London. Clinical Records Interactive Search (CRIS) at South London and Maudsley is a research database based on the SLaM EHR. The pseudonymisation and governance model are explained elsewhere, and form the basis for ethical research that may contain an exploration of free text from records.(Perera, Broadbent et al. 2016) To purpose the records for large-scale studies, natural language processing applications (“apps”) have been designed to extract concepts such as mental health diagnosis, signs and symptoms of illness, and efficacy of medication.(Perera, Broadbent et al. 2016, Mueller, John et al. 2021)

### Medication Prescribing

For studies looking at medication use in dementia, using CRIS or other specialist databases has advantages regarding the clinical features of dementia. However, most clinicians in SLaM and other specialist clinics in the community do not prescribe directly to the patient. Instead, they will advise the GP of any recommended changes, to allow GPs to consider the medication regimen as a whole and ensure consistent prescription after discharge from the specialist clinic. Records of medication in CRIS are therefore usually second-hand lists in free text notes or within letters to the GP, rather than structured fields such as records of electronic prescribing that will be found in primary care EHR. A recent query of SLaM-CRIS data showed that 77% of all medication mentions in people with dementia were in free text. (Mueller, John et al. 2021) The CRIS medication app extracts medication names that appear to be referring to medication usage in the patient from both structured and unstructured text using an internal gazetteer based on the British National Formulary enhanced for possible variants in spelling and other features of recording.

The main use of the CRIS medication app has been for psychotropic medications.(Mueller, Perera et al. 2017, Mueller, John et al. 2021), and most of the enhancement with variant spellings has been concentrated here. It is not yet known how well the extraction of physical health medication can classify patients based on their medication use. For instance, in a planned evaluation of the benefits and harms of the use of preventative medications in people with dementia, the medication app will need to classify aspirin and statin use in the year that dementia is diagnosed. The presence of the link between SLaM-CRIS and a primary care database that covers approximately one-quarter of the SLaM population allows a comparison between prescribing records from primary care and the medication use extracted from a specialist care EHR by the SLaM-CRIS medication app.(Davis, Mueller et al. 2021)

## Methods

### Data Sources

Lambeth DataNet (LDN) is a primary care electronic health record database that captures patient information from Lambeth GPs (formerly Lambeth Clinical Commissioning Group, now part of the SE London Integrated Care Board). Clinical Records Interactive Search (CRIS) is a framework for electronic health record databases, from which we accessed information from the South London and Maudsley NHS Foundation Trust (SLaM-CRIS), which includes dementia care for people from Lambeth. The SLaM-CRIS/LDN linkage is conducted by the CRIS data-linkage service (Perera, Broadbent et al. 2016).

CRIS, including linkage to Lambeth DataNet, has received ethical approval as an anonymized data resource (Oxford Research Ethics Committee C, reference 18/SC/0372). This project was approved by the CRIS oversight committee.

### Cohort

The cohort of patients with dementia diagnoses was defined by ICD-10 diagnosis in SLaM-CRIS and Read code from Lambeth DataNet (which was dual coded Read and SNOMED so that the same list could be run pre- and post-transition to SNOMED), as explored in an earlier paper.(Davis, Mueller et al. 2021) The date of diagnosis, or index date, was the first in either record. From the cohort we excluded any patients who were aged below 65 at date of diagnosis. To ensure that we had the fullest record possible we only selected those patients who were registered with a Lambeth GP at the date of first diagnosis and who had used SLaM services at some time.

Cohort descriptors were derived from structured fields in CRIS and LDN as previously done (Davis, Mueller et al. 2021). Age and sex from LDN; dementia subtype from most recent from CRIS ICD-10 if present, or from LDN Read codes; dementia severity from CRIS using MMSE score (mild > 22, moderate 19-22, severe <19). Comorbidities were probed using Read code lists in the LDN journal at dates up to the index date.

### Choosing medications

The medication classes were statins and aspirin, as per the main project, and eight other preventative medications (see box 1). Preventative medications were suggested by the NICE Multimorbidity research questions: taken long-term to prevent morbidity or mortality, but benefit unclear for people with multimorbidity.(National Guideline Centre 2016) Dementia-specific medications of acetylcholinesterase inhibitors and memantine made an eleventh category. Medication lists based on the classes were made using the British National Formulary.

#### Box 1

Medications (short names in parentheses)

1. Study medications Aspirin Statins
2. Other long-term preventative medications ACE inhibitors and angiotensin receptor blockers (ACEi or ARB) Anticoagulants Beta-blockers Bisphosphonates Calcium with vitamin D (Ca vitD) Non-aspirin antiplatelets Proton pump inhibitors SSRI or mirtazapine antidepressants (antidepressants)
3. Dementia-specific medication

Acetylcholinesterase inhibitors or memantine (dementia drugs)

### Defining medications

The target was to extract the medications that patients were using in the year they were diagnosed with dementia. The *index date* is the date of first dementia diagnosis documentation, with the *index year* starting on that day.

In LDN, the patients’ prescription issue table was searched in the index year to one year later for any medication from the medication list, and classified into the 11 classes. A single prescription issue was classed as usage in the year.

In SLaM-CRIS, the medication app was used to search the record from six months before the index date until six months after the index date, to try to capture the assessment associated with the diagnosis. The medication lists were used alongside the internal gazetteer.

### Secondary analysis (subcohort)

All patients in this sample had at least one episode in SLaM, however, not all patients will have had a full assessment around the time of dementia diagnosis. For instance, someone seen by a liaison psychiatry or a care home may not have a full assessment documented in the SLaM EHR. Those referred to the memory clinic are more likely to have had a thorough assessment documented, including letters to the referrer, which will increase the material available for the medication app.We hypothesised that this group would have better agreement.

### Analysis

Raw numbers were compared, and converted to prevalence estimates for each medication in each database. Confidence intervals at 95% were calculated by Wilson’ s method and differences in estimates between databases presented. To allow comparison between medications of different prevalence, agreement of medication status was calculated using Cohen’ s kappa (agreement adjusted for chance agreement) with 95% confidence intervals. The sensitivity of CRIS to identify prescribing in LDN was defined as the proportion of patients with prescription in LDN who were also identified by CRIS. Specificity is not calculated, since it implies that mentions found in SLaM-CRIS that are not in LDN are in error, which is unlikely to be the case. Medication classes were compared against statin as our reference medication class.

## Results

### Population

4410 people diagnosed with dementia aged 65 and over who had data from both primary care (LDN) and SLaM services (CRIS). Female patients outnumbered male (59% vs 41%), and 63% were aged 80 or over at the time of diagnosis. Dementia in Alzheimer’ s disease was the most common subtype but there was a significant amount of missing data around the dementia subtype (21.2%) and stage of dementia at diagnosis (30.5%). Most patients (2776, 63%) had contact with their GP surgery 24 or more times in the two years prior to dementia diagnosis, equivalent to once per month. Comorbidity is common in this cohort before the index date, with a median Charlson comorbidity score of 2, excluding dementia, and probable diabetes in 34%, history of a stroke in 20% and documented osteoporosis in 13%.

### Prevalence estimates

Table 1 shows the numbers of mentions of each medication class ascertained through the specialist data (CRIS) medication app and through the primary care (LDN) prescription issue table for 4410 people diagnosed with dementia. These are converted to prevalence estimates with 95% confidence intervals based on CRIS mentions and LDN prescription.

**Table 1.**
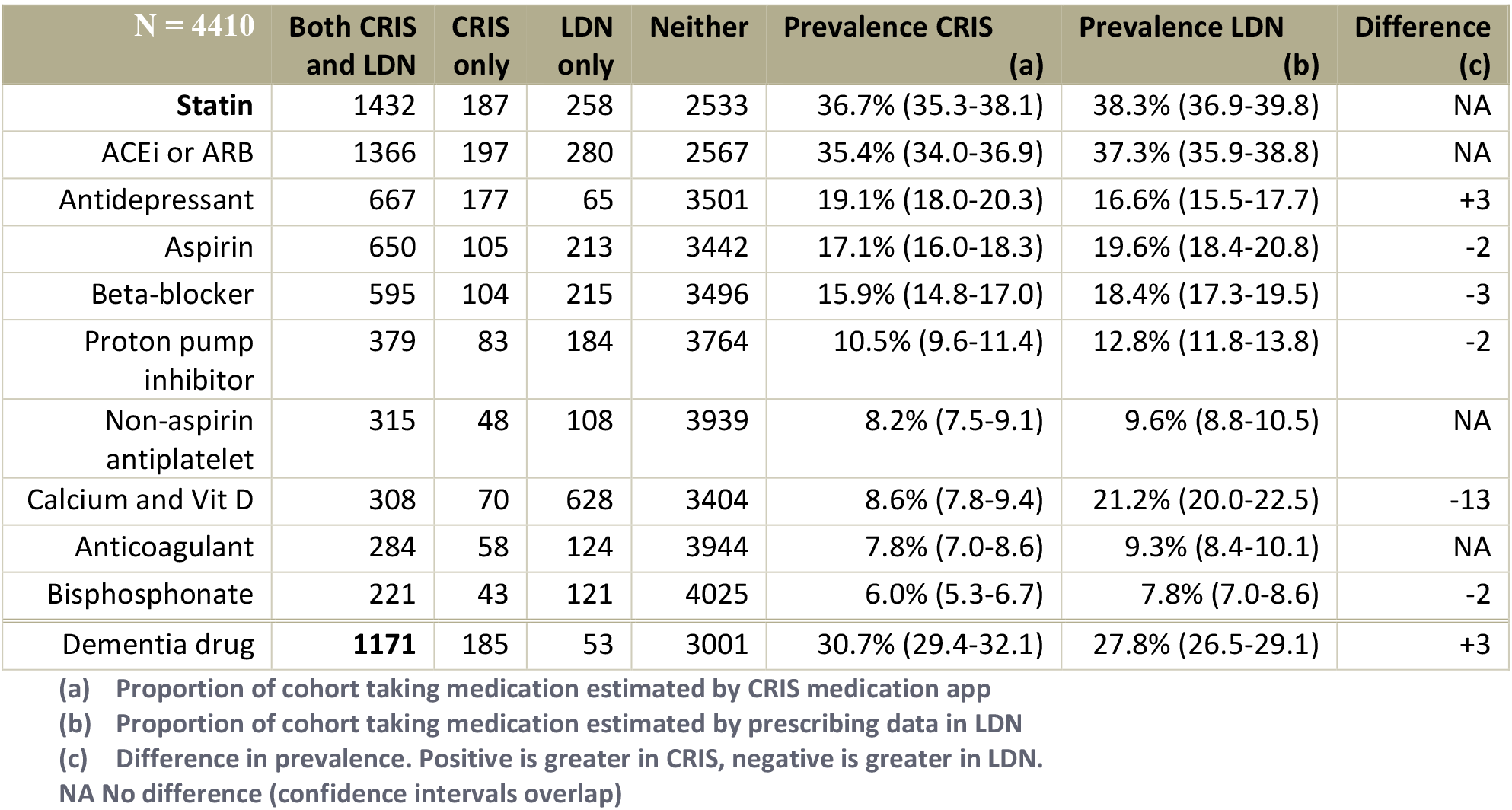
Ascertainment of selected medication use by two methods: CRIS medication app and LDN prescription issue.

| N = 4410 | Both CRIS<br>and LDN | CRIS<br>only | LDN<br>only | Neither | Prevalence CRIS<br>(a) | Prevalence LDN<br>(b) | Difference<br>(c) |
| --- | --- | --- | --- | --- | --- | --- | --- |
| **Statin** | 1432 | 187 | 258 | 2533 | 36.7% (35.3-38.1) | 38.3% (36.9-39.8) | NA |
| ACEi or ARB | 1366 | 197 | 280 | 2567 | 35.4% (34.0-36.9) | 37.3% (35.9-38.8) | NA |
| Antidepressant | 667 | 177 | 65 | 3501 | 19.1% (18.0-20.3) | 16.6% (15.5-17.7) | +3 |
| Aspirin | 650 | 105 | 213 | 3442 | 17.1% (16.0-18.3) | 19.6% (18.4-20.8) | -2 |
| Beta-blocker | 595 | 104 | 215 | 3496 | 15.9% (14.8-17.0) | 18.4% (17.3-19.5) | -3 |
| Proton pump<br>inhibitor | 379 | 83 | 184 | 3764 | 10.5% (9.6-11.4) | 12.8% (11.8-13.8) | -2 |
| Non-aspirin<br>antiplatelet | 315 | 48 | 108 | 3939 | 8.2% (7.5-9.1) | 9.6% (8.8-10.5) | NA |
| Calcium and Vit D | 308 | 70 | 628 | 3404 | 8.6% (7.8-9.4) | 21.2% (20.0-22.5) | -13 |
| Anticoagulant | 284 | 58 | 124 | 3944 | 7.8% (7.0-8.6) | 9.3% (8.4-10.1) | NA |
| Bisphosphonate | 221 | 43 | 121 | 4025 | 6.0% (5.3-6.7) | 7.8% (7.0-8.6) | -2 |
| Dementia drug | **1171** | 185 | 53 | 3001 | 30.7% (29.4-32.1) | 27.8% (26.5-29.1) | +3 |

(a) Proportion of cohort taking medication estimated by CRIS medication app
(b) Proportion of cohort taking medication estimated by prescribing data in LDN
(c) Difference in prevalence. Positive is greater in CRIS, negative is greater in LDN.
NA No difference (confidence intervals overlap)

Figure 2 illustrates that for statins, ACE inhibitors, non-aspirin antiplatelets and anticoagulants, the CRIS prevalence estimate overlapped with the LDN prevalence estimate. For antidepressants and dementia drugs, using CRIS to estimate prevalence would find 3 extra people taking these medications per 100 studied. Ascertainment of aspirin, beta-blockers, proton pump inhibitors and bisphosphonates would be lower by two to three people per 100 from CRIS. The biggest difference is in Ca vitD, which would have a prevalence of 8.6% in CRIS but 21.2% in LDN.

**Figure 1.**
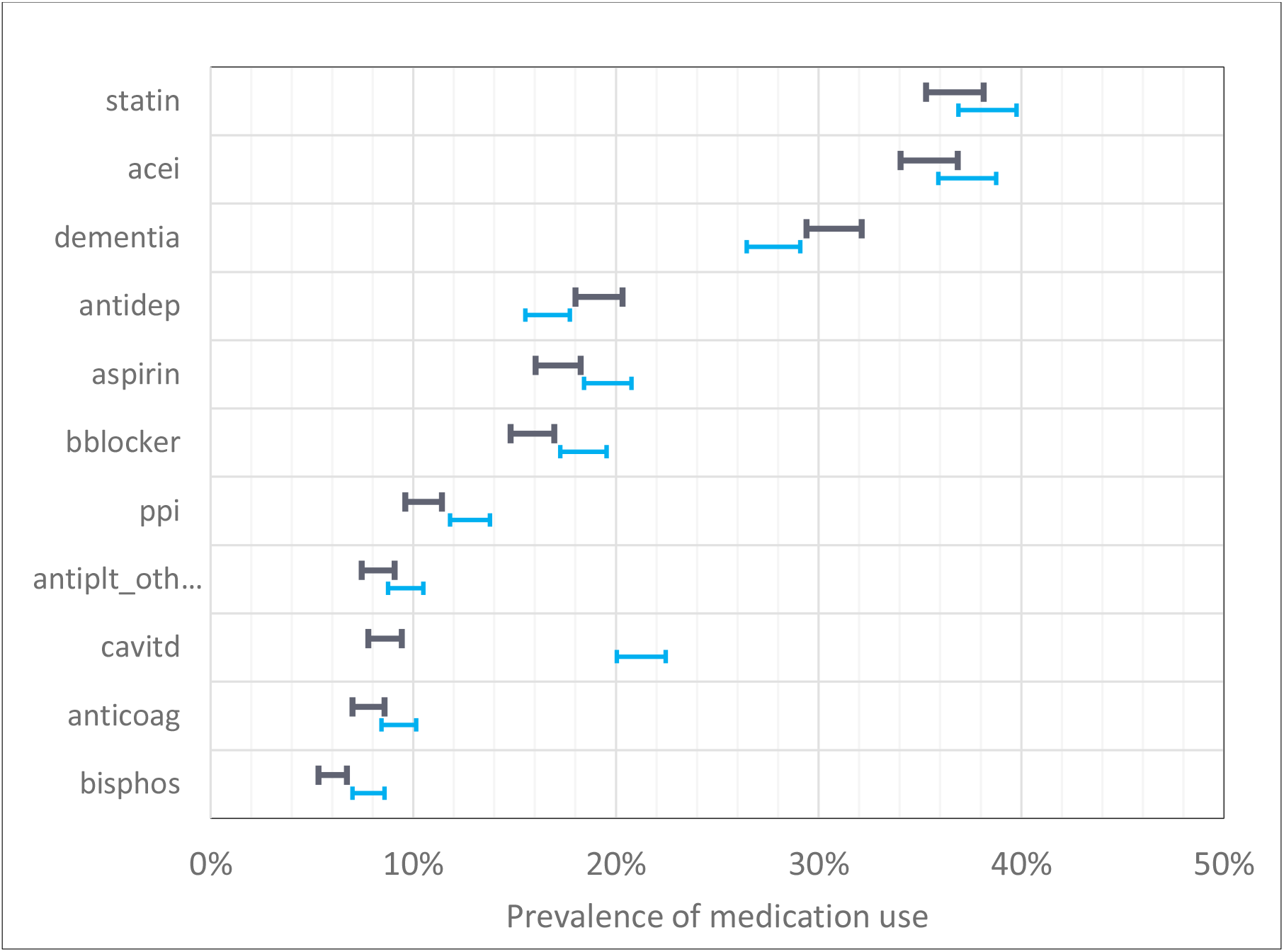
Prevalence of medication in CRIS (black, upper) and LDN (blue, lower). Bars show 95% confidence intervals.

**Figure 2.**
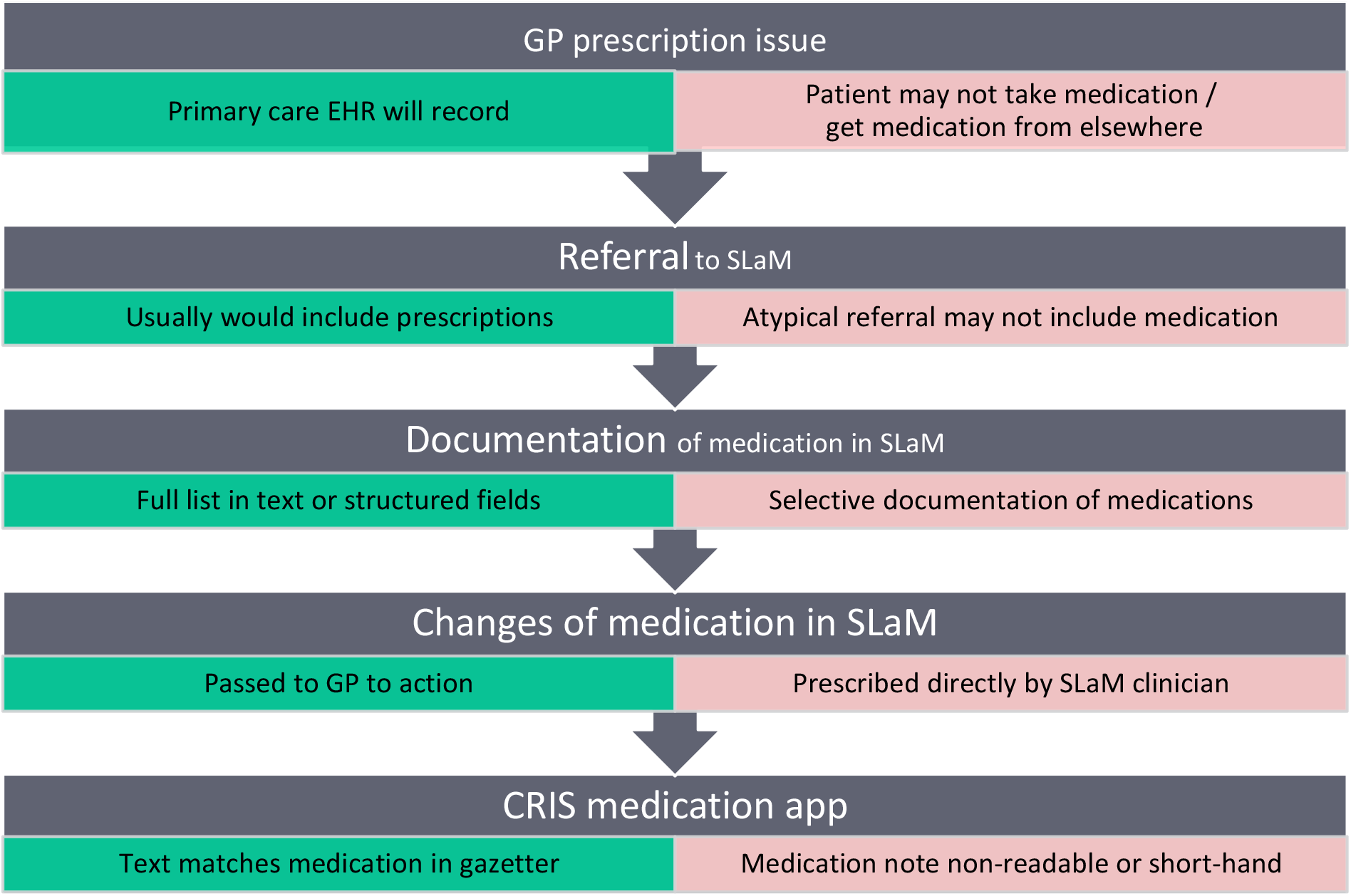
Steps from prescription to ascertainment in CRIS necessary for accurate ascertainment, alongside facilitators (green) and barriers (red) to correct ascertainment.

### Agreement

Comparing the medication status in individuals, agreement (Cohen’ s kappa) between the two methods of ascertainment was above 0.8 for antidepressants and dementia drugs, and above 0.7 for all medication classes except for Ca vitD (at 0.385). Using statins as the reference, antidepressants and dementia drugs had better agreement, and proton pump inhibitors and Ca vitD had worse agreement.

### Sensitivity

Of the people with a statin prescription in LDN, 85% are found by CRIS (sensitivity), as shown in table 2, with other medications taken for the vascular system having a similar sensitivity (70-85%). For psychotropic medications, CRIS had higher sensitivity (96% dementia, 91% antidepressant). For proton pump inhibitors and medications for osteoporosis, it had lower sensitivity (67% proton pump inhibitor, 65% bisphosphonates, 33% for Ca vitD).

**Table 2.**
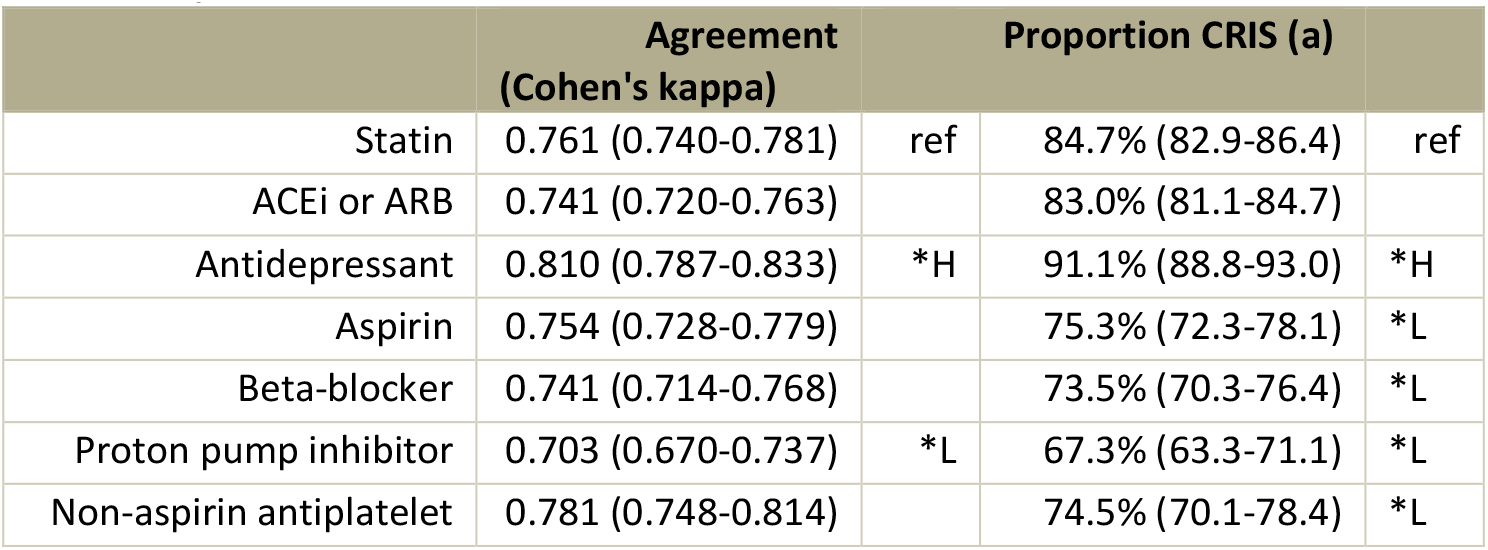
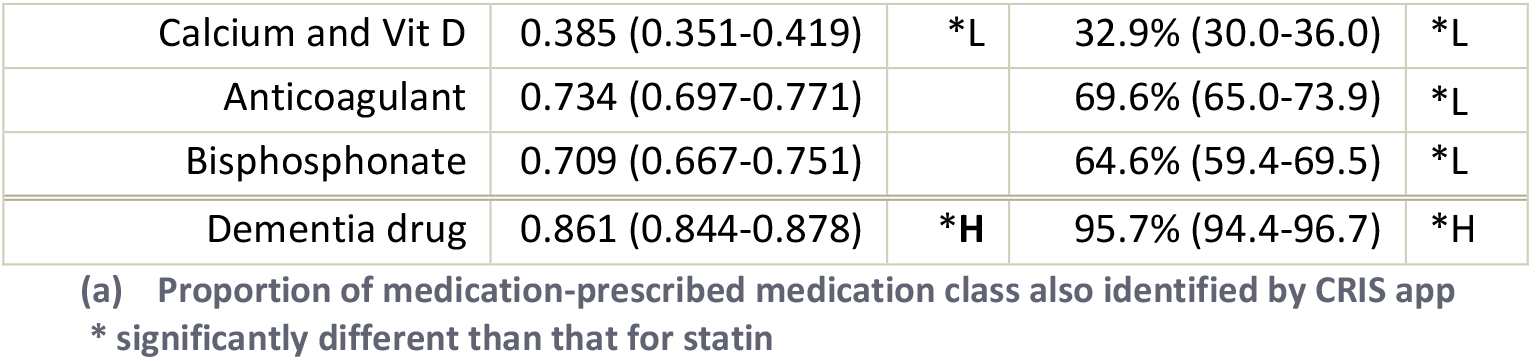
Comparisons between methods to ascertain medication.

|  | Agreement<br>(Cohen's kappa) |  | Proportion CRIS (a) |  |
| --- | --- | --- | --- | --- |
| Statin | 0.761 (0.740-0.781) | ref | 84.7% (82.9-86.4) | ref |
| ACEi or ARB | 0.741 (0.720-0.763) |  | 83.0% (81.1-84.7) |  |
| Antidepressant | 0.810 (0.787-0.833) | *H | 91.1% (88.8-93.0) | *H |
| Aspirin | 0.754 (0.728-0.779) |  | 75.3% (72.3-78.1) | *L |
| Beta-blocker | 0.741 (0.714-0.768) |  | 73.5% (70.3-76.4) | *L |
| Proton pump inhibitor | 0.703 (0.670-0.737) | *L | 67.3% (63.3-71.1) | *L |
| Non-aspirin antiplatelet | 0.781 (0.748-0.814) |  | 74.5% (70.1-78.4) | *L |
| Calcium and Vit D | 0.385 (0.351-0.419) | *L | 32.9% (30.0-36.0) | *L |
| Anticoagulant | 0.734 (0.697-0.771) |  | 69.6% (65.0-73.9) | *L |
| Bisphosphonate | 0.709 (0.667-0.751) |  | 64.6% (59.4-69.5) | *L |
| Dementia drug | 0.861 (0.844-0.878) | *H | 95.7% (94.4-96.7) | *H |

(a) Proportion of medication-prescribed medication class also identified by CRIS app
\* significantly different than that for statin

### Secondary analysis

Limiting to the 2103 people referred to the memory clinic did not substantially alter the patterns of ascertainment (table 3).

**Table 3.** Comparison of ascertainment of medication from two sources (CRIS medication app and LDN prescription), the subgroup of those referred to the memory clinic n=2103.

|  | Agreement (Cohen's kappa) | Proportion CRIS (a) |
| --- | --- | --- |
| Statin | 0.744 (0.715-0.773) | 86.1% (84.0-88.0) |
| ACEi or ARB | 0.716 (0.686-0.746) | 84.6% (82.4-86.7) |
| Antidepressant | 0.801 (0.766-0.836) | 88.1% (84.1-91.1) |
| Aspirin | 0.762 (0.728-0.795) | 78.2% (74.4-81.7) |
| Beta-blocker | 0.748 (0.714-0.783) | 74.8% (70.9-78.4) |
| Proton pump inhibitor | 0.710 (0.667-0.753) | 68.7% (63.6-73.4) |
| Non-aspirin antiplatelet | 0.786 (0.743-0.829) | 75.1% (69.4-80.1) |
| Calcium and Vit D | 0.370 (0.326-0.414) | 33.8% (30.1-37.7) |
| Anticoagulant | 0.740 (0.694-0.787) | 68.7% (62.9-74.0) |
| Bisphosphonate | 0.718 (0.665-0.771) | 68.3% (61.7-74.2) |
| Dementia drug | **0.837 (0.812-0.862)** | **95.6% (94.1-96.8)** |

(a) Proportion of medication-prescribed medication class also identified by CRIS medication app

## Discussion

In a cohort of 4410 people diagnosed with dementia in old age between 2009 and 2019, a linked dataset allowed a comparison between prescribing data from primary care (LDN) and medication extracted from a specialist EHR database (SLaM-CRIS). Medications taken long-term for prevention, plus medication taken for dementia were studied, with prescribing in the index year in LDN compared against medication mention in SLaM-CRIS around the time of dementia diagnosis.

Agreement (kappa) in all but one class was above 0.7, with Ca vitD being lower at 0.385. The ability of the specialist data to identify prescription in the window was over 90% for psychotropics and between 70-90% for cardiovascular medications, but lower for proton pump inhibitors, bisphosphonates, and Ca vitD. We discuss possible reasons for these results in “authors’ interpretation” below.

For the specific question of how well the CRIS data can select those prescribed aspirin and statin in the index year, this analysis suggests that CRIS SLaM can detect 85% of patients taking statins and 75% of those taking aspirin. The higher sensitivity for statin is probably related to the higher prevalence of statin use, as kappa agreement (which corrects for chance agreement) finds them much more similar (0.761 (0.740-0.781) for statin, 0.754 (0.728-0.779) for aspirin). Such levels of agreement are usually said to be ‘ good’, (Landis and Koch 1977) and since some of the different classifications may be informative, this data is likely to be adequate for research use with appropriate caution.

## Authors’ interpretation

A discussion with clinicians and others related to these databases revealed several different barriers to the correct ascertainment of patient medication, as shown in Box 2 that were related to a pathway shown in figure 2.

### Box 2

Issues contributing to patient medications differing from what documented. Read in conjunction with figure 2

**Box 2 Issues contributing to patient medications differing from what documented. Read in conjunction with figure 2**
| Issue | Point in pathway | Effect on ascertainment | Other comments |
| --- | --- | --- | --- |
| **Patients may not take all medication prescribed** | GP prescription issue | LDN would over-ascertain. CRIS may not, depending on whether patient was candid with SLaM clinicians | We think medications such as Ca vitD and bisphosphonates that are difficult to take may sometimes fall into this category |
| **Patients may take medication from elsewhere** | GP prescription issue / referral / documentation | LDN would under-ascertain. CRIS may not, depending on what the patient told the SLaM clinician | Ca vitD, proton pump inhibitors, statins and aspirin are available from pharmacists or over the counter |
| **SLaM-clinician (SC) does not have record of prescribing** | Referral | CRIS will under-ascertain | This should be becoming less as EHRs are better connected |
| **SC has many medications to document, and may decide to omit some** | Documentation | CRIS will under-ascertain medications that are not directly related to mental state / dementia | This would apply especially to PPIs and medications for osteoporosis |
| **SC may not document, or document in a way not readable (e.g. saying “see referral”)** | Documentation / CRIS medication app | CRIS will under-ascertain | Electronic referral may be readable by the app |
| **SC may prescribe medication for patient to try, rather than asking GP to prescribe** | Changes of medication in SLaM | LDN would under-ascertain, but assuming that prescribing is eventually passed to GP, this would only be temporary | This would apply to psychotropics in SLaM, with a parallel in every clinic the patient attends or every time in hospital. |
| **SC may document some medications in short-hand** | CRIS medication app | CRIS will under-ascertain these medications, but app could be recalibrated to improve detection | This applies especially for supplements used as medication, eg Ca vitD |

Prescribing data from Lambeth DataNet in the index year will be a highly accurate report of the prescribing by Lambeth NHS GPs. It will not be able to represent prescribing elsewhere, for example by hospitals or if patients move away during the year. In particular, for this study, there is the possibility of medication being prescribed by SLaM. Medication changes suggested by SLaM clinicians are usually passed to the GP to implement – for dementia drugs, this often involves a shared care agreement.(National Institute for Health and Care Excellence 2018, South East London Area Prescribing Committee 2019) However, SLaM clinicians can prescribe themselves where appropriate. Since the prevalence of psychotropic medication is higher when extracted from SLaM-CRIS, these may have been prescribed by SLaM clinicians and not continued by the GP (i.e. if it were tried and stopped due to adverse effects).

A prescription issue does not always mean that a patient will have taken the medication, and there is not currently any way of ascertaining this by routinely collected data. One may suspect that a patient who is not taking their medication as prescribed would order them less frequently than indicated, but our analysis considered one prescription in the year to be positive. A future study might utilise the data to look at the number of doses ordered over a year to look at intermittent versus routine use. Some misclassification in the specialist data may be reflecting true differences in what is prescribed by Lambeth GP and what is taken.

Recording of medication by clinicians in the SLaM will be standard during assessments and reviews. It may be recorded in the medication tab, in a ‘ progress note’ or an attachment (such as a letter to the GP). Medication will be taken from the referral, via a link to the shared care record (that will include primary care info) or reported by the patient. This copying of medication will be prone to error and to the clinician selectively recording according to relevance in the assessment. Historically referrals may have been paper only as a letter or fax, more recently they were scanned into the EHR as a picture that cannot be ‘ read’ by the medication app. Electronic referrals, attached to the EHR as text documents, will offer advantages for natural language processing. Selective report may account for the high sensitivity of the specialist data for psychotropics, and relatively less for bone-related medications. Compounding this, errors in spelling or non-standard ways of recording medications may be missed by the CRIS medication NLP app. This may explain the very low sensitivity of Ca vitD: for instance, “calcium with calciferol” or “Adcal D3” will be detected as Ca vitD, whereas “calcium” or “Adcal” would not.

One more reason for the less-than-perfect agreement is the slightly different time windows used for different databases, so that something prescribed late in the index year in primary care may not be captured by specialist care in the six months index date. However, these windows were used to answer the specific question for the research project, regarding prescribing around the time of dementia diagnosis, and not an audit of the process of recording.

### Strengths and weaknesses

This innovative research linkage of data from primary care to a specialist dementia care provider allows a fuller and more complete picture of the health of people with dementia. This is done securely with pseudonymised notes and a governance system that involves patients from SLaM. Results will go not only into research projects but will be fed back to improve the tools available in CRIS. However, the exclusion of patients without SLaM records and any patients are opted out of registry-related research nationally or locally, may provide some loss of representativeness.

The limitations in the gazetteer, which is mainly trained on psychotropic medication may lead to under-ascertainment of physical health medications, as we found, but work is ongoing to rectify this, and these improvements may mean that the results published here are surpassed in subsequent versions. Specifically to this study, the periods of observation were not completely aligned, which may also underestimate the potential of the medication app.

### Implications

These results indicate that SLaM-CRIS data on medication around the time of a dementia diagnosis is not a perfect reflection of prescribing in primary care, but for common medications and psychotropics it shows high sensitivity and good agreement. Using the CRIS medication app is common in studies using SLaM-CRIS, but results should be critically assessed for the potential for physical health medication to be under-recorded.

CRIS is used for other mental health trusts, who use similar NLP apps. The results described here may be reassuring to users of CRIS, but as we discuss in “authors’ interpretation”, the results represent the outcome of GP, patient and clinicians from the mental health trust, so there may be some differences in other times and places that cannot be readily predicted. For users of other databases, these results may not be generalisable, but the over challenge and influences discussed may help to inform the use of medication mentions into their studies.

## Supporting information

STROBE checklist

## Data Availability

Data produced in the present study may be protected by the data model of SLaM-CRIS and LDN. Please contact the SLaM-CRIS administrator for details of how to apply for data or call the CRIS team on 0203 228 8553

## Acknowledgements

With thanks to the teams at LDN and CRIS for supporting this study, particularly Amelia Jewel at the CRIS data linkage service and Mark Ashworth who commented on a version of this manuscript.

